# General anesthesia dissociates discrete components of ketamine neurophysiology

**DOI:** 10.1101/2025.08.05.25333019

**Authors:** Ben Deverett, Duan Li, Theresa Lii, Phillip E. Vlisides, Vijay Tarnal, Anna Forsyth, Rachael Sumner, Pilleriin Sikka, Alan F. Schatzberg, Suresh Muthukumaraswamy, George A. Mashour, Boris D. Heifets

## Abstract

**Importance:** Ketamine has well known dissociative, analgesic, and antidepressant properties, but it is unknown whether the neurophysiologic effects that are associated with these properties can be modulated separately from one another. Given prior studies that link specific cortical oscillations with specific therapeutic effects, it is likely that modulating selective aspects of ketamine neurophysiology can inform efforts to develop more targeted therapies.

**Objective:** To determine how the neurophysiologic signatures of ketamine are influenced by removal of conscious awareness using general anesthesia.

**Design:** Observational cohort study from trials spanning 2017 to 2023.

**Setting:** Multicenter study using data from two study cohorts (U-Michigan and Stanford) and supplementary analysis of a third cohort (U-Auckland).

**Participants:** 52 participants in primary analysis and 27 additional participants in supplementary analyses. Study cohorts included healthy volunteers, patients undergoing elective surgery, and patients with a diagnosis of depression.

**Intervention:** Participants received a subanesthetic infusion of ketamine (0.5 mg/kg) or placebo with or without general anesthesia (GA).

**Main Outcome and Measure:** Changes in electroencephalographic (EEG) band power during medication infusion.

**Results:** GA differentially alters EEG features commonly associated with ketamine. In comparison to awake administration, ketamine during GA preserves its high-frequency power modulation but lacks its characteristic low-frequency augmentation.

**Conclusions and Relevance:** Co-administration of ketamine with GA selectively modulates the high- and low-frequency neurophysiological correlates of ketamine, suggesting a method to explore these components’ role in ketamine’s behavioral effects.

## Introduction

Ketamine is a widely used anesthetic, analgesic, and antidepressant^1,2^. Low doses produce connected consciousness with dissociative experiences and high doses produce a state of disconnected dissociative anesthesia. These subanesthetic and anesthetic states are characterized by elevations in βγ (high-frequency) and θ (low-frequency) power on electroencephalography (EEG)^3–9^. Ketamine’s antidepressant mechanisms have been attributed to high-frequency oscillations^10^ while its dissociative properties have been linked to low-frequency oscillations^5,11^. However, there are no known methods to independently modulate these aspects of ketamine-induced neurophysiology in humans. Separating these mechanisms can inform efforts to develop ketamine-like antidepressant therapies without inducing dissociation.

We hypothesized that removal of conscious awareness, and therefore removal of dissociative experience, could be used to study which features of ketamine neurophysiology are related to experience. We predicted that ketamine administered under general anesthesia (GA; induced by non-dissociative GABAergic agents) would exhibit different EEG features compared to ketamine administered awake. Preclinical studies suggest that anesthetics disrupt ketamine-evoked neurotransmission in frontal regions^12^ and its associated γ oscillations^10,13,14^, and θ oscillations have been associated with subjective experiences of ketamine dissociation^11^. We therefore hypothesized that ketamine administered under GA would lack these high- and low-frequency oscillatory modulations. We compared EEG recorded from patients receiving ketamine under GA or receiving ketamine only.

## Methods

We analyzed data from two studies utilizing a standardized ketamine infusion: 0.5 milligrams per kilogram body weight over 40 minutes. In Study 1 (U-Michigan)^4,15^, 15 healthy participants received ketamine in a laboratory session (“ketamine-only”). In Study 2 (Stanford)^16^, from which EEG data have not been published, 37 patients under steady-state GA for elective surgery were administered ketamine (n=18) or placebo (n=19) intraoperatively (“ketamine-GA” and “placebo-GA” respectively) while anesthetized using standard regimens (Table S1). Data from an additional Study 3, (U-Auckland)^17,18^ in which 27 participants with depression were administered ketamine, were used to assess the impact of depression on the EEG effects of ketamine.

Study 1 data were collected using a 128-channel HydroCel Geodesic sensor, Study 2 data were collected intraoperatively using the SedLine frontal EEG monitor, and Study 3 data were collected using a 64-channel BrainCap MR. Data were preprocessed with downsampling, filtering, and artifact removal as described in the original studies and Supplementary Methods. To compare frontal EEG across datasets, channels corresponding to the SedLine electrode locations (Fp1=L1, Fp2=R1, F7=L2, F8=R2) were extracted and re-referenced to Fz, and all subsequent analyses were performed in an identical manner across datasets. We separately confirmed the consistency of EEG cap and SedLine data using simultaneous recordings from the same individual, which showed comparable spectral profiles (Fig. S1).

Spectrograms were computed using the multitaper method described in Supplementary Methods. For spectral analyses in Fig. 1, spectral power was extracted individually for each participant in the bands θ (theta, 4-8 Hz), Lβ (low-beta, 15-22 Hz), and βγ (beta-gamma, 23-40 Hz) using the median power within the band. Fold-Δ power was computed as above on a per-subject basis then averaged across subjects for display. Decibel power values were displayed in the positive range using a constant additive offset applied uniformly to all datasets.

**Figure 1:**
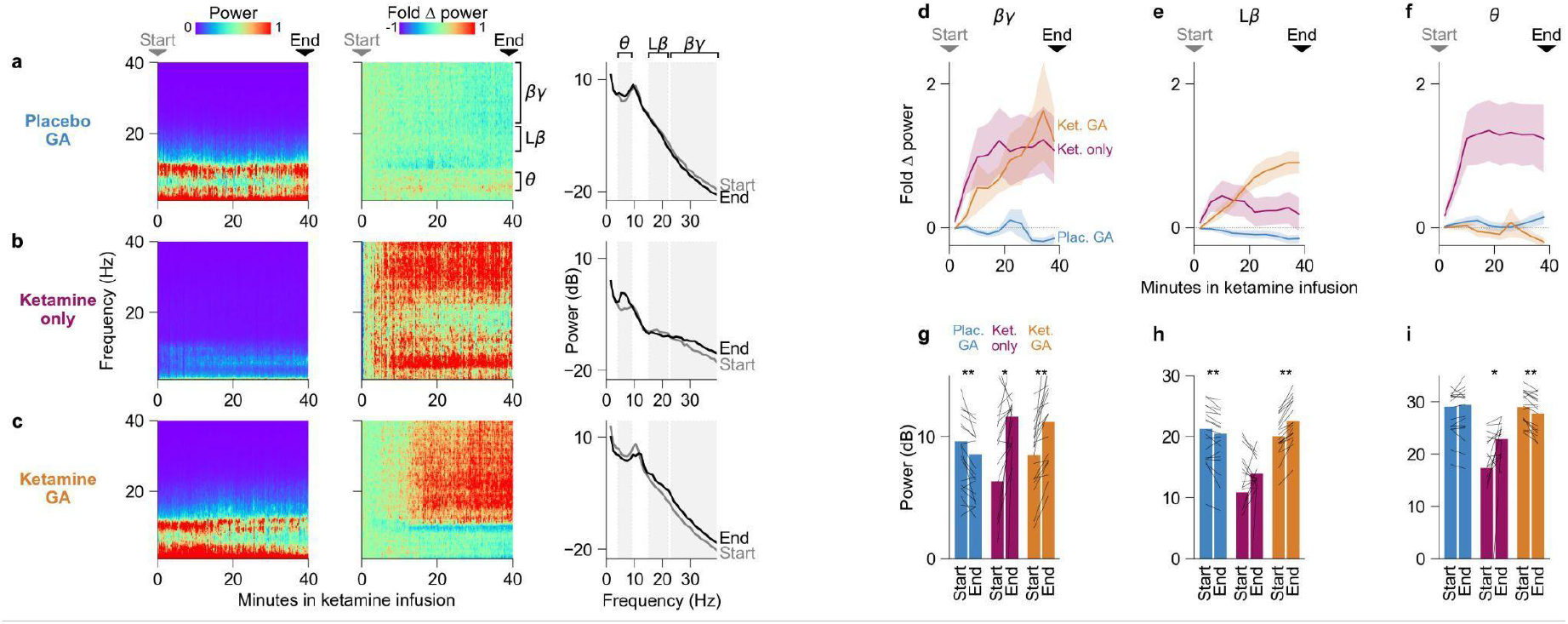
EEG spectral profiles during ketamine/placebo infusions. (a-c) Left: spectral power time courses for placebo-GA (n=19), ketamine-only (n=15), and ketamine-GA (n=18), median across participants, normalized from 0 to 1. Middle: fold spectral power change relative to Start of infusion, normalized from -1 to 1. Fold Δ power is computed as (p-p0)/p0 where p0 is the baseline power in the first 3 minutes of infusion on a per-subject and per-frequency basis. Right: power spectral density at the Start and End of infusion, median across participants. Start and End times correspond to the median values within the 3 first minutes and final minutes of the infusion, respectively. **(d-f)** Time course of fold power change relative to Start of infusion, mean ± SEM over participants, in three frequency bands (left: βγ, middle: Lβ, right: θ). **(g-i)** Mean power at Start and End of infusion for all experimental conditions and frequency bands. Lines denote individual participants. ^*^: p<0.05; ^**^: p<0.01; two-tailed Wilcoxon signed-rank test.

## Results

Under GA-only, patients exhibited stereotypical anesthetized EEG profiles (Figure 1a) characterized by α (8-14 Hz) and Δ (0.5-4 Hz) oscillations. In contrast, the ketamine-only group exhibited typical signatures of subanesthetic ketamine: increases in βγ and θ power^3,4^(Fig. 1b, middle; Fig. 1d,f), rising within 10 minutes and remaining elevated throughout the infusion.

Contrary to our hypothesis, the ketamine-GA group showed robust βγ enhancement (Fig. 1c, 1d). A moderate early enhancement of Lβ occurred in both ketamine-only and ketamine-GA but persisted in a significant manner by the end of infusion only in the ketamine-GA group (Fig. 1e). Finally, as predicted, in the ketamine-GA group we observed a total absence of the θ modulation seen with subanesthetic ketamine infusions (Fig. 1f). To quantify these effects, we compared absolute power values across the start and end of the infusion. The following spectral power changes were observed (Fig. 1g-i): in βγ, the placebo-GA group exhibited a decrease (9.6 ± 5.5 vs 8.5 ± 5.3 dB), while the ketamine-only and ketamine-GA groups exhibited increases (6.3 ± 11.3 vs 11.6 ± 2.2 dB and 8.5 ± 2.9 vs 11.2 ± 3.8 dB, respectively). In Lβ, placebo-GA exhibited a decrease (21.4 ± 6.4 vs 20.5 ± 6.5 dB), ketamine-only exhibited an upward but non-significant trend (10.9 ± 11.3 vs 14.0 ± 2.3 dB), and ketamine-GA exhibited an increase (20.1 ± 3.2 vs 22.6 ± 3.5 dB). Finally, in θ, placebo-GA showed no change (29.2 ± 6.3 vs 29.5 ± 6.7 dB), ketamine-only exhibited an increase (17.3 ± 10.5 vs 22.9 ± 3.1 dB), and ketamine-GA exhibited a small decrease (29.0 ± 3.0 vs 27.8 ± 3.5 dB). High-frequency modulation could be an artifact of ketamine-induced EMG activity even under anesthesia, however 27 of 37 anesthetized patients had received a neuromuscular blocking drug within an hour of ketamine infusion.

To address whether the diagnosis of depression in Study 2 could account for these results, we compared data from Study 3 (U-Auckland)^18^ in which patients with depression were administered a subanesthetic bolus of ketamine without GA. These EEG data recapitulated key features of our findings, demonstrating that subanesthetic ketamine causes a rise in βγ and θ power even in the presence of depression (Fig. S2).

## Discussion

We demonstrate that ketamine delivered during GA preserves the enhancement of frontal high-frequency but not low-frequency power, compared to ketamine delivered when awake. The absence of low-frequency modulation on a background of GA was predictable, as dissociative experiences associated with ketamine are presumably absent under GABA-mediated unconsciousness. Together these findings raise the possibility that the ketamine-GA EEG signature reflects the absence of subjective experience (absence of θ increase), alongside the maintenance of other biological effects (persistence of βγ increase). This interpretation is supported by several studies demonstrating that ketamine retains antidepressant efficacy when delivered during surgical anesthesia^16,19–21^, though we note an exception from a secondary analysis of patients receiving ketamine to prevent postoperative delirium^22^. We therefore propose that some, but not all, of ketamine’s biological activity is preserved under GA, and that the mechanisms underlying ketamine’s effects may be separable with targeted perturbations.

Depression was present in the two GA groups but not in the ketamine-only group, however patients with depression exhibit clear θ and βγ modulation (Fig. S2) demonstrating that depression cannot account for the present results. Surgery involving other concomitant medications was also occurring in the two GA groups but not in the ketamine-only group, but these factors do not explain the altered response to ketamine since previous findings in surgical settings where ketamine was delivered without GA demonstrated the same increases in low- and high-frequency power observed in the ketamine-only group of this study^3^. We therefore interpret the observed differences between the ketamine-only and ketamine-GA groups as resulting from the GABA-mediated unconsciousness rather than from differences in patient population.

In summary, we demonstrate that the typical EEG signature of ketamine (elevated βγ and θ power) is selectively altered when ketamine is administered under general anesthesia, preserving high-frequency (βγ) modulations while lacking low-frequency (θ) augmentation. This result suggests that the neuronal mechanisms underlying ketamine’s effects can be selectively altered, which may give way to more precise characterization and usage of ketamine.

## Data Availability

All data produced in the present study are available upon reasonable request to the authors

## Acknowledgements

This work was supported by a grant awarded to B.D.H. by the Society for Neuroscience in Anesthesia and Critical Care. T.R.L. received salary support through a T32 grant from the NIH National Institute on Drug Abuse (3T32DA035165-02S1). B.D. received support from the Research in Anesthesia Training Program (ReAP, NIH T32GM089626). The U-Michigan study was funded by NIH grant R01GM111293. The funding bodies supporting this study had no influence on the conduct of the trial, analysis of the data, or reporting of the results.

## Supplementary Materials

### Supplementary Methods

Analyses were performed in Python 3.12.1 using numpy 1.26.4, scipy 1.11.4, and pandas 2.2.3. In the Stanford cohort, the SedLine device was a Masimo Root v2.1.2.4 with SedLine firmware 2343, and data were exported in EDF format at the conclusion of each surgery. EEG data were collected for 37 of the 40 subjects in the original study (the remaining three had no EEG data collected). Raw data for each patient consisted of four frontal EEG traces sampled at 178 Hz. For four datasets, signal sampling rate and amplitude were adjusted prior to analyses to account for variations in device data output. The two edge channels on the device output frequently exceeded the dynamic range of the measurement device and exhibited clipping, so we excluded these channels from analyses. For each patient, the remaining channels were averaged. Data were bandpass filtered between 0.1 and 50 Hz with a Butterworth filter of order three and notch filtered at 60 Hz. We applied a custom artifact rejection algorithm which excluded data points whose amplitude deviated significantly from the baseline distribution: for each subject we excluded any contiguous 1-second period during which greater than 10% of data points had amplitudes that differed by greater than 2.5 standard deviations from the mean of the entire dataset. Additionally, we excluded any contiguous 100-millisecond period during which greater than 5% of data points had amplitudes that differed by greater than 4 standard deviations from a 3-second rolling average of the dataset. This resulted in 4.2 ± 2% (mean ± s.d.) of samples rejected from further analyses. Data from the U-Michigan and U-Auckland studies were processed as described in the original publications using standard downsampling, filtering, and artifact rejection. To compute spectrograms, data from raw preprocessed time series were processed using the multitaper method^23^ with a non-overlapping window size of 15 seconds, frequency resolution 1 Hz, time half bandwidth 7.5, and 14 tapers.

**Supplementary Figure 1:**
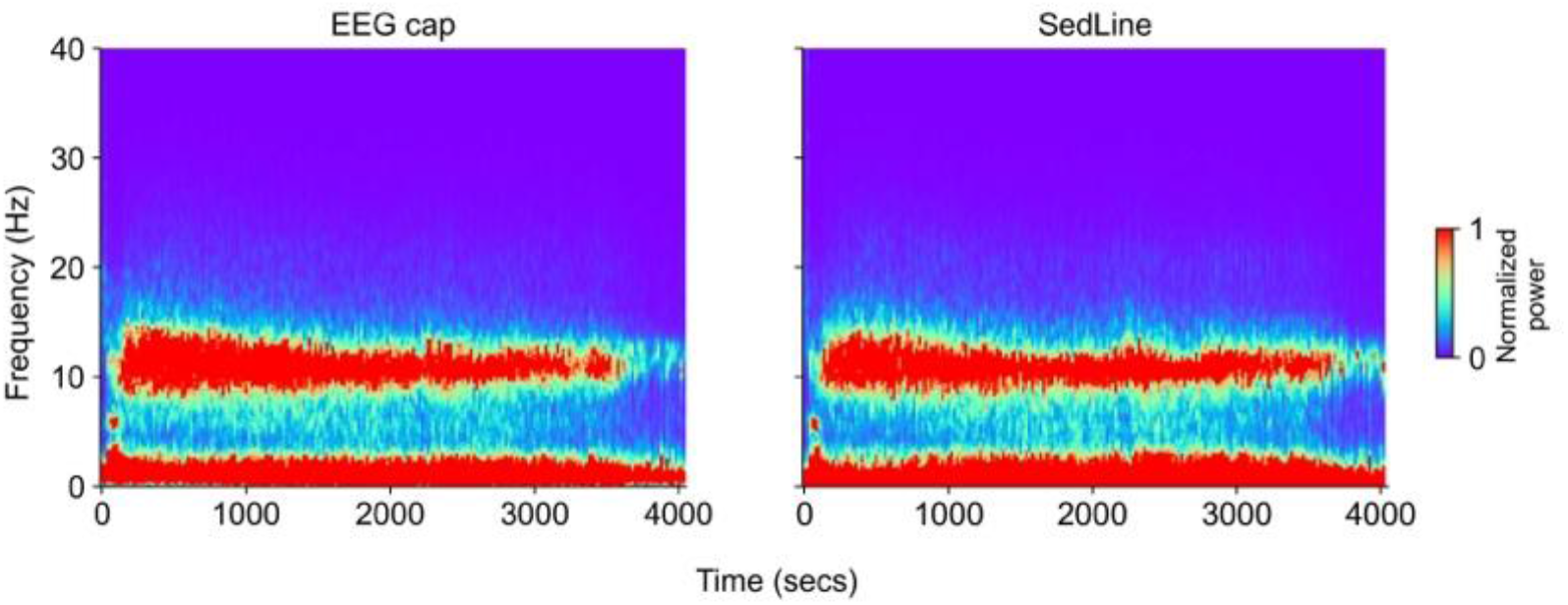
EEG data obtained from SedLine are comparable to those obtained from high-channel EEG cap. These data were captured during a separate experiment involving propofol infusions in healthy participants and are displayed as a general comparison of cap data and SedLine data. The left panel shows data obtained from a 32-channel BrainVision BrainCap. The right panel shows data obtained from the SedLine device. The two signals were simultaneously recorded and processed in the same manner as the present study.

**Supplementary Figure 2:**
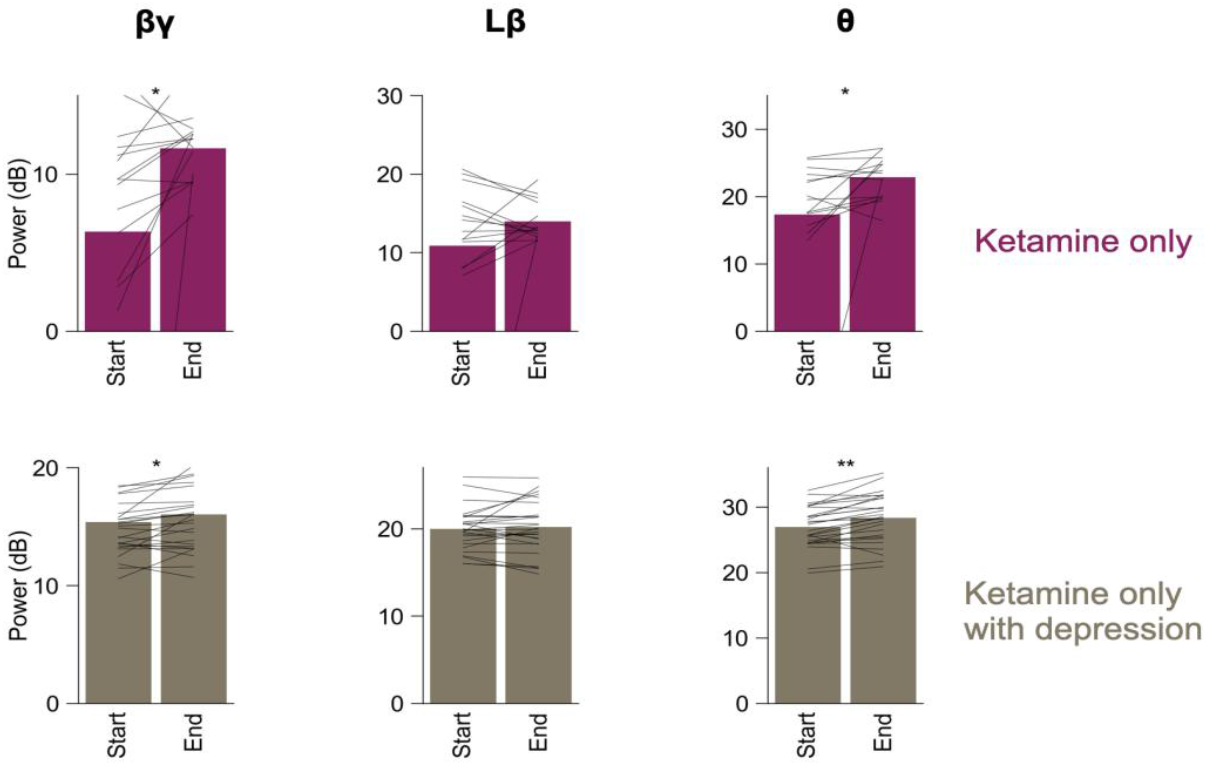
Subjects with and without major depression exhibit the same EEG trends with subanesthetic ketamine. Top row: ketamine-only data from the U-Michigan study as shown in the analyses in Figure 1. Bottom row: data from a separate study (U-Auckland) of 27 participants with major depression receiving a 0.25 mg/kg ketamine bolus. Data were analyzed at time zero (Start) and at 2 minutes after ketamine administration (End), which for a 70-kilogram 40-year-old person corresponds to a ketamine effect-site concentration of 0.2 mcg/ml; this is comparable to the level reached at the End time point of the 0.5 mg/kg infusions analyzed in this study (0.2-0.21 mcg/ml). Conventions are as in Fig. 1g-i.

**Supplementary Table 1:** Maintenance general anesthesia agents and dosages used during ketamine infusions.

| Subject medication | Propofol (mcg/kg/min) | Sevoflurane (end tidal %) | Isoflurane (end tidal %) | Nitrous oxide (end tidal %) | Dexmedetomidine (mcg/kg/hr) | Remifentanyl (mcg/kg/min) |
| --- | --- | --- | --- | --- | --- | --- |
| placebo | 50 | 1.1-1.2 | 0 | 0 | 0 | 0 |
| ketamine | 75 | 0.6-1.6 | 0 | 0 | 0 | 0 |
| placebo | 75 | 0 | 0.2-0.3 | 0 | 0 | 0.05-0.1 |
| placebo | 0 | 0 | 0.3-0.5 | 50 | 0 | 0 |
| ketamine | 75 | 0.8-1 | 0 | 0 | 0 | 0 |
| ketamine | 125 | 0 | 0 | 0 | 0 | 0 |
| placebo | 0 | 1.3-1.7 | 0 | 0 | 0 | 0 |
| ketamine | 50 | 0.8-1.1 | 0 | 0 | 0 | 0.05-0.1 |
| placebo | 30-50 | 1.1-1.4 | 0 | 0 | 0 | 0 |
| ketamine | 0 | 0.6-1.1 | 0 | 50 | 0 | 0 |
| ketamine | 0 | 0.4 | 0 | 55-71 | 0 | 0 |
| placebo | 0 | 0.4 | 0.5-1.1 | 0 | 0 | 0 |
| ketamine | 50-100 | 0.5-0.6 | 0 | 0 | 0 | 0.05-0.15 |
| placebo | 75 | 0.9 | 0 | 0 | 0 | 0 |
| ketamine | 40-50 | 0.6-1 | 0 | 0 | 0 | 0 |
| placebo | 80-125 | 0 | 0 | 0 | 0 | 0.05-0.2 |
| ketamine | 40-90 | 0.9-1.1 | 0 | 0 | 0 | 0 |
| placebo | 50 | 1-1.6 | 0 | 0 | 0 | 0 |
| ketamine | 35-45 | 0.6-1 | 0 | 0 | 0 | 0 |
| placebo | 50 | 1.1-1.3 | 0 | 0 | 0 | 0 |
| ketamine | 50 | 0.5-1.2 | 0 | 0 | 0.6 | 0 |
| placebo | 75 | 0.9-1 | 0 | 0 | 0 | 0 |
| placebo | 150 | 0 | 0 | 0 | 0 | 0.2 |
| ketamine | 75 | 0 | 0 | 0 | 0 | 0.05-0.1 |
| ketamine | 50 | 1-1.2 | 0 | 0 | 0 | 0 |
| ketamine | 50 | 1.3-1.4 | 0 | 0 | 0 | 0 |
| placebo | 50 | 0.9 | 0 | 0 | 0 | 0 |
| placebo | 0-50 | 0.8-0.9 | 0 | 0 | 0 | 0 |
| placebo | 50 | 1.6-2 | 0 | 0 | 0 | 0 |
| ketamine | 50 | 1-1.1 | 0 | 0 | 0 | 0 |
| ketamine | 50 | 1 | 0 | 0 | 0 | 0 |
| ketamine | 50 | 1.1-1.3 | 0 | 0 | 0 | 0 |
| ketamine | 50 | 0.6-0.8 | 0 | 0 | 0 | 0.1 |
| placebo | 50 | 0.8 | 0 | 0 | 0 | 0 |
| placebo | 95 | 0 | 0 | 0 | 0 | 0 |
| placebo | 0 | 0.9 | 0 | 0 | 0 | 0 |
| placebo | 90-100 | 0 | 0 | 0 | 0 | 0-0.1 |

**Supplementary Table 2:** Participant demographics.

|  | U-Michigan ketamine | Stanford-GA ketamine | Stanford-GA placebo | U-Auckland ketamine |
| --- | --- | --- | --- | --- |
| # of participants | 15 | 18 | 19 | 27 |
| Age, yrs (Mean ± SD) | 24.9 ± 5.8 | 50.0 ± 12.4 | 51.9 ± 19.3 | 30.2 ± 8.2 |
| Sex (M/F) | 7/8 | 7/11 | 4/15 | 12/15 |

